# Forecasting Novel Corona Positive Cases in India using Truncated Information: A Mathematical Approach

**DOI:** 10.1101/2020.04.29.20085175

**Authors:** Brijesh P. Singh

## Abstract

Novel corona virus is declared as pandemic and India is struggling to control this from a massive attack of death and destruction, similar to the other countries like China, Europe, and the United States of America. India reported 2545 cases novel corona confirmed cases as of April 2, 2020 and out of which 191 cases were reported recovered and 72 deaths occurred. The first case of novel corona is reported in India on January 30, 2020. The growth in the initial phase is following exponential. In this study an attempt has been made to model the spread of novel corona infection. For this purpose logistic growth model with minor modification is used and the model is applied on truncated information on novel corona confirmed cases in India. The result is very exiting that till date predicted number of confirmed corona positive cases is very close to observed on. The time of point of inflexion is found in the end of the April, 2020 means after that the increasing growth will start decline and there will be no new case in India by the end of July, 2020.

## Introduction

A novel corona virus is responsible for epidemic popularly known as COVID-19 is a new strain that has not been identified previously in humans. WHO declared COVID-19 a pandemic on March 11, 2020.^[1]^ The virus that caused the incidence of Severe Acute Respiratory Syndrome (SARS) in 2002 in China, Middle East respiratory syndrome (MERS) in 2012 in Saudi Arabia and the virus that causes COVID-19 are genetically related to each other, but the diseases they caused are quite different.^[2]^ These viruses, in general, are a family of viruses that target and affect mammal’s respiratory systems. The SARS corona virus spread to humans via civet cats, while the MERS virus spread via dromedaries. In case of the novel corona virus, typically happens via contact with an infected animal, perhaps the common carriers are bats initial reports from seafood market in central Wuhan, China.

Novel corona virus is spreading throughout the world at alarming speed. Worldwide it has exploded to 1118684 cases and caused 58909 deaths by April 3, 2020.^[3]^ Developed countries like Italy, Spain, France and United State of America etc. are struggling to overcome from the pressure created by novel corona virus. India with a huge population about 1.3 billion, amongst majority of the people are living in poor hygienic condition and the medical facilities like number of doctors and hospitals are less in India as compared to developed countries indicates that the situation of India will become very critical but comparatively better public health system and political control in India than the above developed countries. India reported 2545 cases novel corona confirmed cases as of April 2, 2020 and out of which 191 cases were reported recovered and 72 deaths occurred. The first case of novel corona is reported in India on January 30, 2020 when a student returned from Wuhan, China.^[4]^ The Government of India was quick to launch various levels of travel advisories beginning from February 26, 2020, with restrictions on travel to China and nonessential travel restrictions to Singapore, South Korea, Iran and Italy.^[5]^ The efforts to control by the Hon’ble Prime Minister Narendra Modi Ji through Janata Curfew (public curfew) on March 22, 2020, can be seen as the beginning of wide-scale public preventive measures. India has launched several social distancing measures and personal hygiene measures during the second week of March.^[6]^

Since huge population of about 1.3 billion, thus India has chosen a flexible strategy of large-scale quarantine and limited testing because of less number of testing kits and also the cost of testing is too much. The country is relying on the people power; thousands of health-care workers are working out across the country to trace and quarantine people who might have had contact with those with novel corona. People are typically only tested if they develop symptoms. Countries such as South Korea isolated infected people based on widespread testing, but some scientists say that India’s mass surveillance approach could achieve a similar goal, and be relevant for other low and low-middle income countries facing kit shortages. Under the lockdown, people are allowed out for essentials, such as food and medical care, but in most states people under quarantine are closely monitored by social workers and cannot leave their homes in some places. If public health workers do not trace all infected individuals during the lockdown, India will need to continue its period of stringent physical distancing.

For the spread of novel corona virus, when disease dynamics are still unclear, mathematical modeling helps us to estimate the cumulative number of positive cases in the present scenarios. Now India is interring in the mid stages of the epidemic. It is important to predict how the virus is likely to grow amongst the population. A mathematical modeling approach is a suitable tool to understand the dynamics of epidemic. In the study some mathematical approach to understand the dynamics of novel corona virus in India has been discuss.

## Methodology

We obtained the truncated information on cumulative number of corona positive confirmed cases in India from March 13 to April 2, 2020 from covid19india.org.^[4]^ All cases are laboratory confirmed following the case definition by the Govt. of India. Some studies modeled the epidemic curve obeying the exponential growth.^[7, 8]^ The nonlinear least square framework is adopted for data fitting and parameter estimation for 2019-nCoV at this early stage. In this study first exponential and then logistic growth curve has been used to model the novel corona epidemic, since epidemics grow exponentially not linearly. But it is surprising that exponential growth curve always provide increasing number of daily new cases. There is no saturation point. Another deterministic model used for understanding the dynamics of epidemic is the SIR model, which has been used to accurately predict incidence like SARS. In the SIR model, we need to know the input parameters first the stats we feed into the model.^[9, 10, 11]^ The first one is *R_0_* called the basic reproduction Number. It is essentially the number of new cases a single infected person will cause during their infectious period. It is one of the most important parameters for assessing any epidemic. Corona virus has an *R_0_*~2.4. In contrast, the H_1_N_1_ virus had an *R_0_*~1.5 in the 2009 swine flu epidemic.^[12]^ The *R_0_* will inform us about how many people will get infected with one infected person. Other one is the case fatality rate (CFR), which is the percentage of infected people that will die due to the infection. The CFR for corona virus has been reported between 0.5–4%. The lower values are more appropriate in resource better settings of medical facility. But SIR model assumes that every person is moving and has equal chance of contact with each and every other person among the population irrespective of the space or distance between different people. It is assumed that the transmission rate remains constant throughout the period of pandemic. Also this model considered to have the same transmission rate for who have been diagnosed and are in quarantine or those who have not been quarantined. The harmonic analysis methods and dynamic model estimates show that the number of COVID-19 infected would be 9225 (if there were 10 infected individuals as of March 1, 2020, who was not taking any precautions to spread), 17,986 (if there were 20) and 44,265 (if there were 50).^[13]^

## Growth Models

A growth curve is an empirical model of the evolution of a quantity over time. Growth curves are widely used in biology for quantities such as population size in population ecology and demography for population growth analysis, individual body height in physiology for growth analysis of individuals. Growth is also a key property of many systems such as an economic expansion, spread of an epidemic, the formation of a crystal, an adolescent’s growth and the condensation of a stellar mass.

### Linear growth

This is the simplest growth model, in which population grows at a constant rate over time. Linear growth is described by the equation

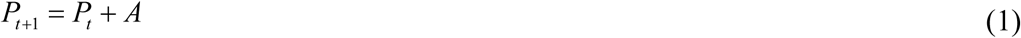

where *P_t_* represents the numbers or size of the system at time *t*, *P_t_*_+1_ represents the system’s numbers or size of the system one time unit later, and *A* is the system’s (linear) growth rate. Many times this model fails to explain natural phenomenon.

*Exponential growth* (Unlimited population growth)

Another simple model describes exponential growth, in which population grows at a constant proportional rate over time. The relation may be expressed in either of two forms, depending on whether reproduction is assumed to be continuous or periodic.^[14]^ Exponential growth results in a continuous curve of increase or decrease, whose slope varies in direct relation to the size of the population.

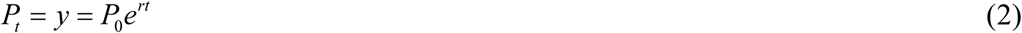

where *r* is the constant rate of growth, *P_o_* is the initial population size, and the variables *t* and *P_t_* respectively represent time and the population at time *t* (Method 1). Another form of exponential curve is as follows

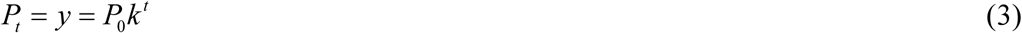

Where 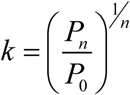 and that therefore the growth rate in (3) does not a constant growth rate. David A. Swanson, University of California, USA used this type equation for prediction (Method 2). We have used truncated information i.e. only 30 days information (from March 4 to April 2, 2020) on number of corona confirmed cases for the prediction purpose. We have used two equations of exponential curve given below

I. 28*e*^014^*^t^* up to March 31, 2020
II. 28*e*^015^*^t^* from March 31, 2020 onward (adjusting faster rate of occurrence of corona cases due to Tablighi spread)

With the current incidence of the novel corona virus going on, we hear about exponential growth. In this study, an attempt has been made to understand and analyze the data through exponential growth curve. The reason for using exponential growth curve for studying the pattern of novel corona virus incidence is that epidemiologists have studied these types of happenings and it is well known that the first period of an epidemic follows exponential growth. The exponential growth function is not necessarily the perfect representation of the epidemic. I have tried to fit exponential curve first, and at the next point to study the logistic growth curve because exponential curve is only fit the epidemic at the beginning. At some point, recovered people will not spread the virus anymore and when someone is or has been infected, the growth will stop. Logistic Growth is characterized by increasing growth in the beginning period, but a decreasing growth after point of inflexion. For example in the corona virus case, the maximum limit would be the total number of exposed people in India because when everybody is infected, the growth will be stopped. After that the increasing rate of curve starts to decline and reach to the minimum.

In the figure 1, predicted values of the cumulative number of novel corona positive cases obtained by method 1 and 2 is drawn along with observed cumulative number of novel corona positive cases. Bothe the methods provide moderately good estimates but the tendency of both the curves are unlimited increasing. The rate of growth of Method 2 is slightly lesser than the rate of growth of Method 1. The number of total infected cases by April 30, 2020 would be about 144700 (Method 1) and 127700 (Method 2). If we do not adjust the Method 1 for Tablighi spread then the total infected cases by April 30, 2020 would be about 81810. Thus we can obtain the effect of Tablighi spread is about 75 percent.

**Figure 1.**
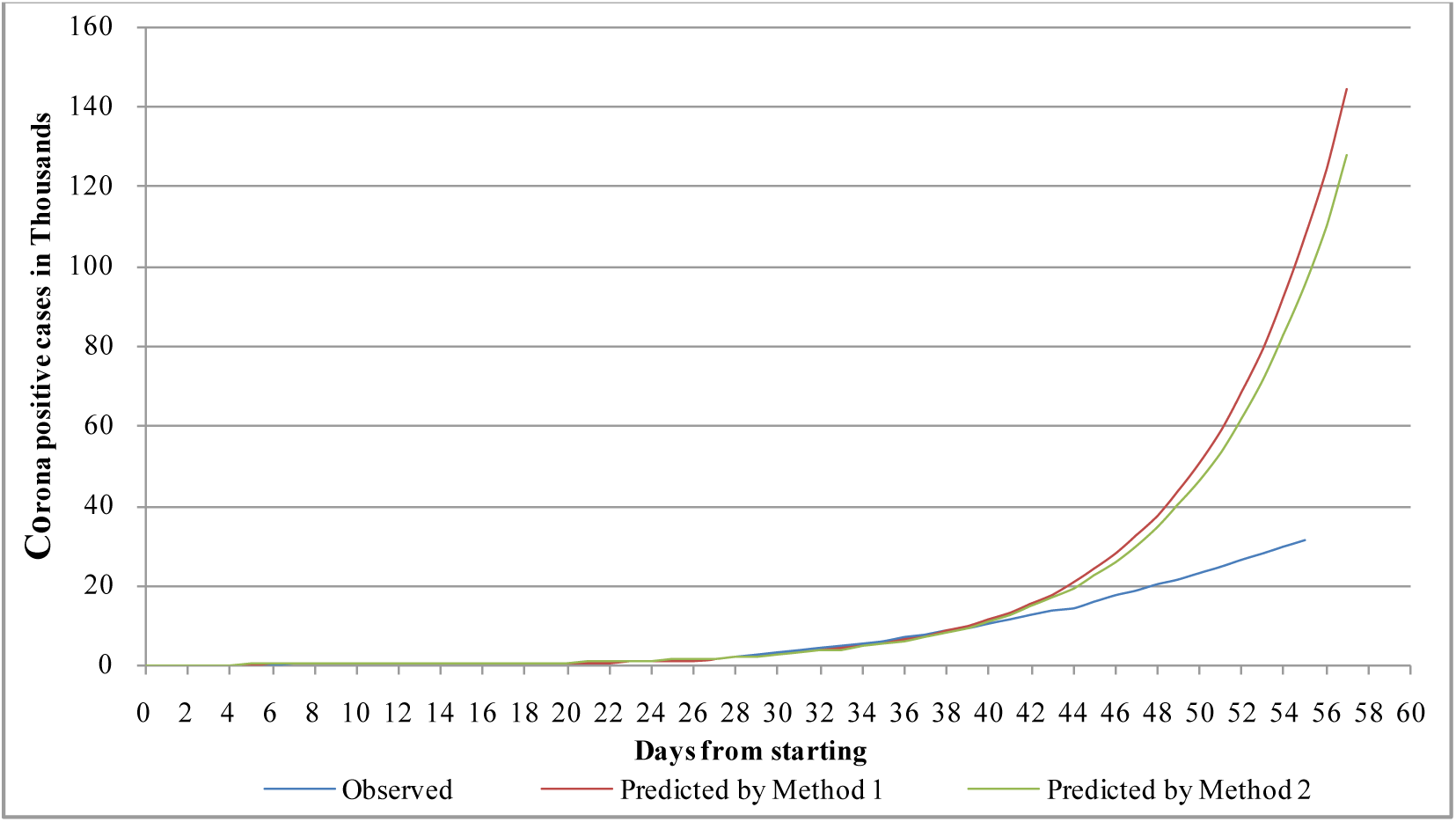

### *Logistic growth* (Sigmoidal)

The logistic model reveals that the growth rate of the population is determined by its biotic potential and the size of the population as modified by the natural resistance, or, in other words, by all the various effects of inherent characteristics, that are density dependence.^[15]^ Natural resistance increases as population size gets closer to the carrying capacity. Logistic growth is similar to exponential growth except that it assumes an essential sustainable maximum point. In exponential growth curve, the rate of growth of *y* per unit of time is directly proportional to *y* but in practice the rate of growth cannot be in the same proportion always. The logistic curve will continue up to certain level, called the level of saturation, sometimes called the carrying capacity, after reaching carrying capacity it starts declining. The factor *y* is called the momentum factor which increases with time *t* and the factor *(k* − *y)* is known as the retarding factor which decreases with time. A system far below its carrying capacity will at first grow almost exponentially however, this growth gradually slows as the system expands, finally bringing it to a halt specifically at the carrying capacity.^[14, 15]^ The logistic relationship can be expressed as

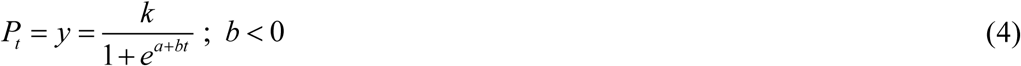

Logistic curve has a point of inflexion at half of the carrying capacity *k*. This point is the critical point from where the increasing rate of curve starts to decline. The time of point of inflexion can be estimate as 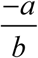. For the estimation of parameter of logistic curve, method of three selected point has been used. The estimate of the parameters can be obtained with equation given as:

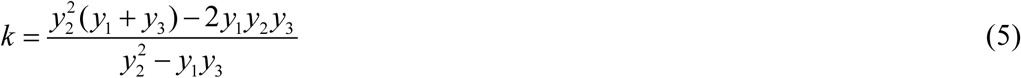

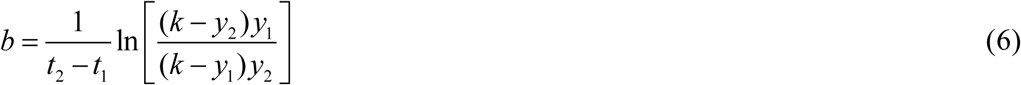

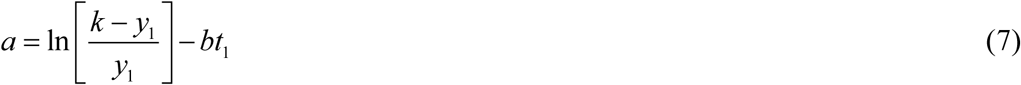

Where *y*_1_, *y*_2_ and *y*_3_ are the number at given time *t*_1_, *t*_2_ and *t*_3_ respectively provided that *t*_2_ − *t*_1_ = *t*_3_ − *t*_2_. You may also estimate the parameter *a* and *b* by method of least square after fixing *k*.

To predict confirmed corona cases on different day, logistic growth curve has been also used and found very exciting results. The truncated information on confirmed cases in India has been taken from March 13 to April 2, 2020. The estimated value of the parameters are as follows *k*=18708.28, *a*=5.495 and *b*=-0.174, with these estimates predicted values has been obtained and found considerably lower values than what we observed. On April 1 and 2, 2020 the number of confirmed corona cases are drastically increasing in some part of India due to some unavoidable circumstances thus there is an earnest need to increase carrying capacity of the model, thus it is increased and considered as 22000 and the other parameters *a* and *b* are estimated again which are *a*=5.657 and *b*=-0.173. The predicted cumulative number of cases is very close to the observed cumulative number of cases till date. The time of point of inflexion is obtained as 32.65 i.e. 35 days after beginning. We have taken data from March 13, 2020 so that the time of point of inflexion should be April 14, 2020 and by May 30, 2020 there will be no new cases found in the country. The distribution of the new cases is in the red color in the figure 2, which is quite normal and obvious. As mentioned in the above paragraph Method 1 provided natural estimate of the total infected cases by May 30, 2020 is 192400. This estimate is obtained when no preventive measure would be taken by the Government of India. The testing rate is lower in India than many western countries, so our absolute numbers is low, when government initiate faster testing process then we have observed more number of cases and fount this logistic model fail to provide cumulative number of corona confirm cases after April 17, 2020 thus there is a need to modify this model. In order to the modification I have taken natural log of cumulative number of corona confirm cases instead of cumulative number of corona confirm cases as taken in the previous model. This model provides the carrying capacity is about 77000 cases and time of point of inflexion April 30, 2020. The present model provides reasonable estimate of the cumulative number of confirmed cases till date (see Appendix and figure 3) and by the end of July, 2020 there will be no new cases found in the country.

**Figure 2.**
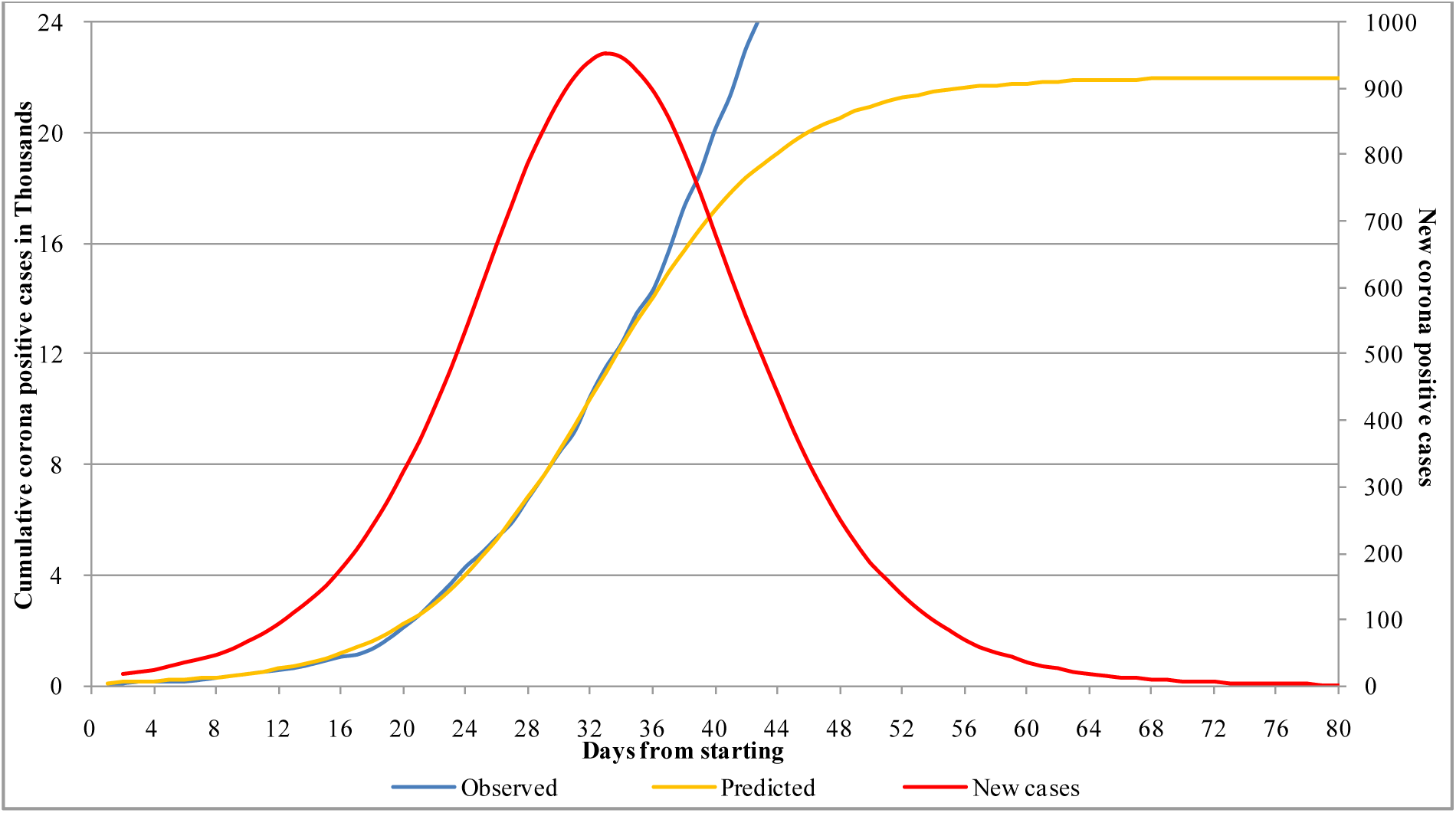

**Figure 3.**
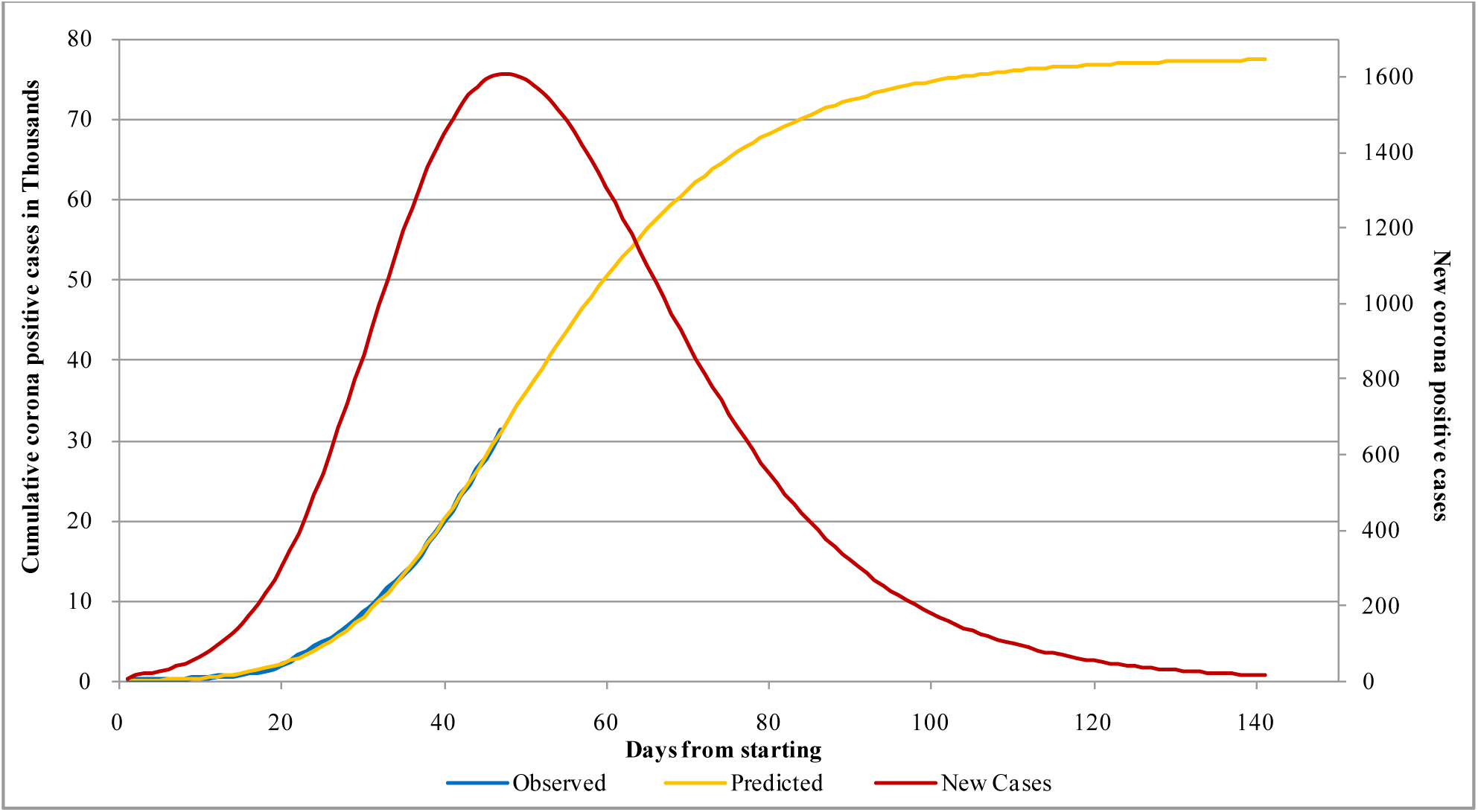

## Conclusions

India is in the comfortable zone with a lower growth rate than other countries studied. Our mathematical model shows that, the epidemic is likely to stabilize with 77000 cases by the end of July, 2020. A study advocated of 49 days single lockdown i.e. lockdown up to May 13, 2020 reduce infected cases below 10 but our study contradict it.^[11]^ The government has adopted a strategy of large scale quarantine and limited testing to flatten the epidemic curve and reduce the death rate. The projections produced by the model and after their validation can be used to determine the scope and scale of measures that government need to initiate. In conclusion, if the current mathematical model results can be validated within the range provided here, then the social distancing and other prevention, treatment policies that the central and various state governments and people are currently implementing should continue until new cases are not seen. The spread from urban to rural and rich to poor populations should be monitor and control is an important point of consideration. Mathematical models have certain limitations that there are many assumptions about homogeneity of population in terms of urban/rural or rich/poor that does not capture variations in population density.

## Data Availability

given in the appendix of the paper

---

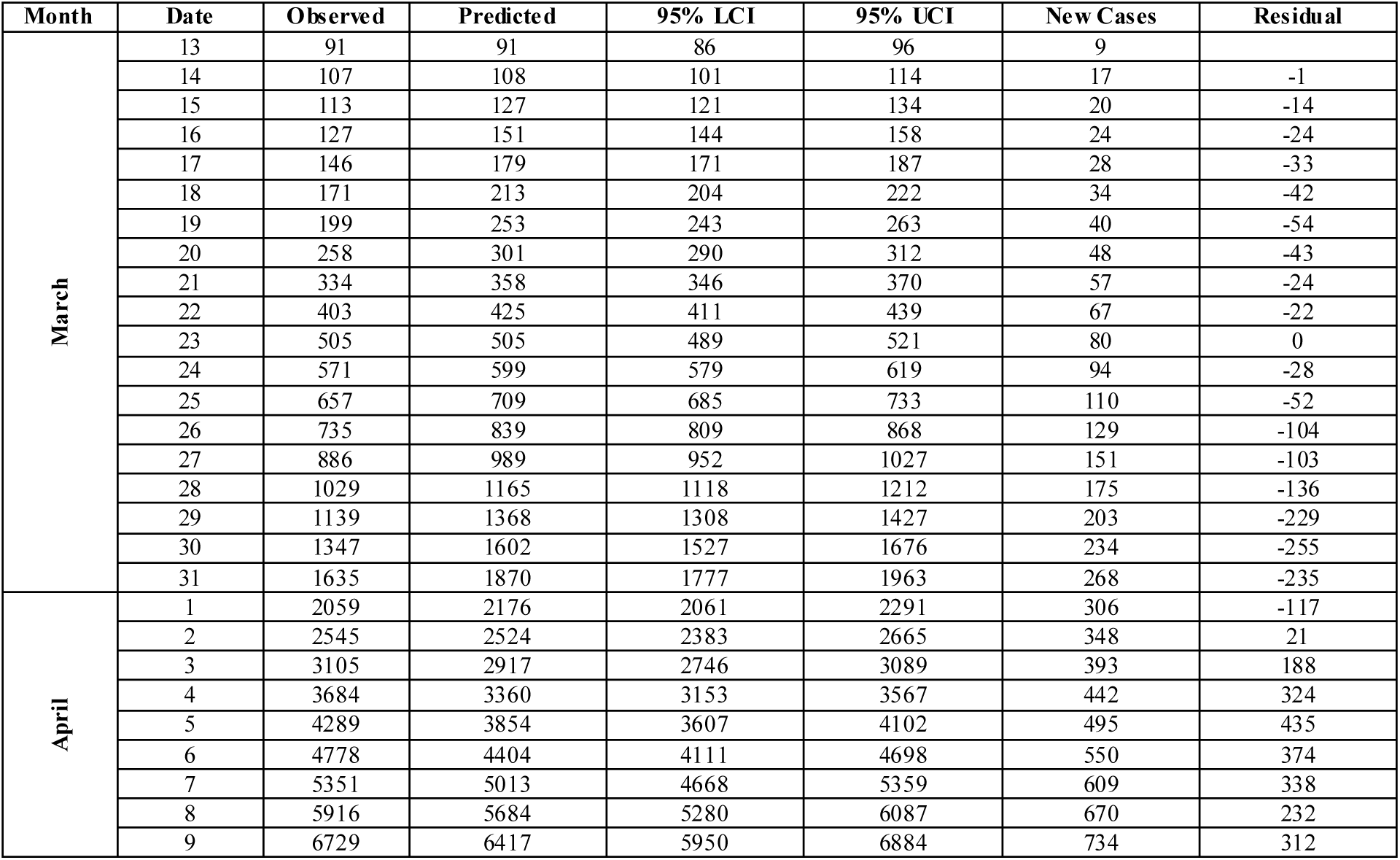
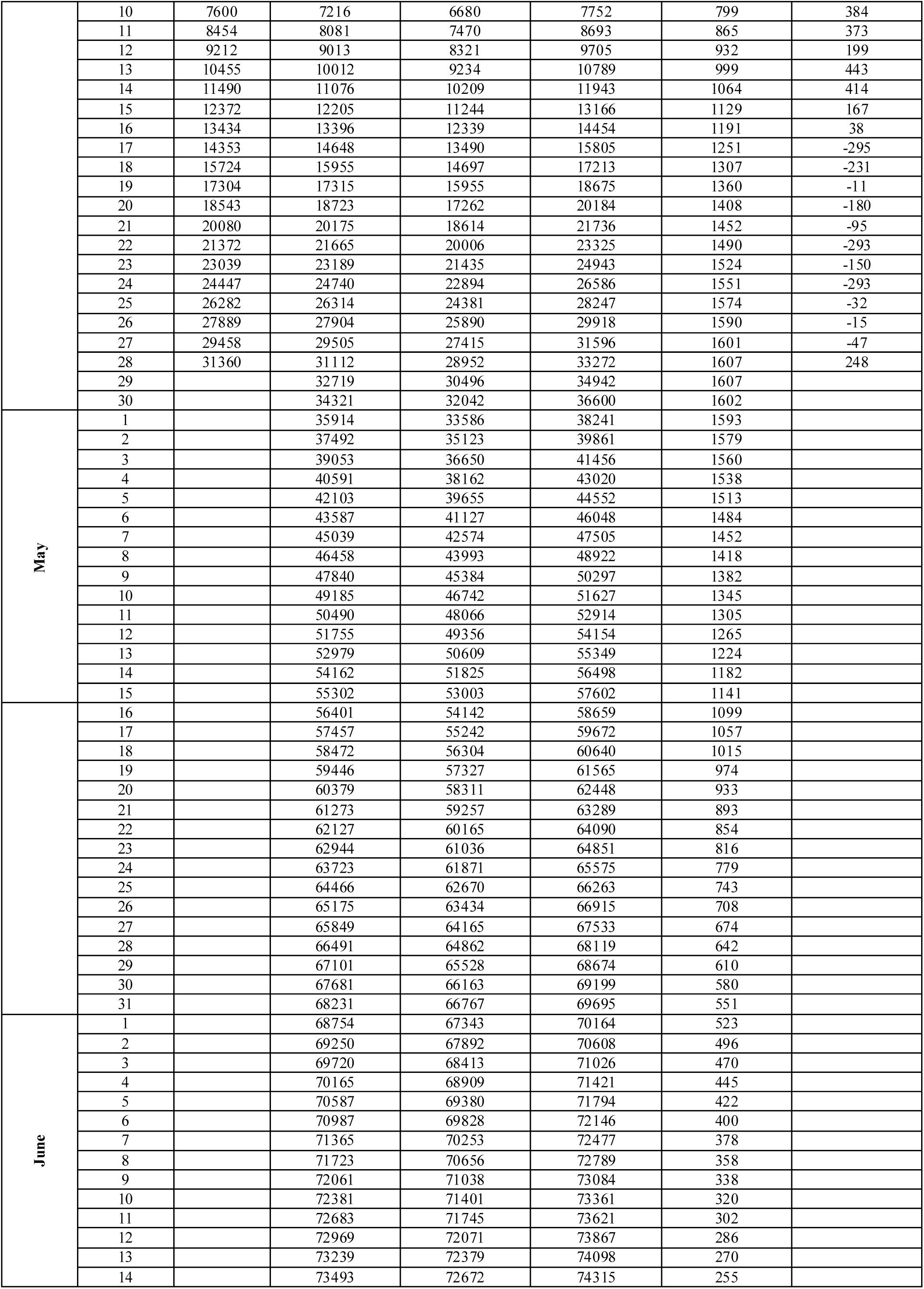
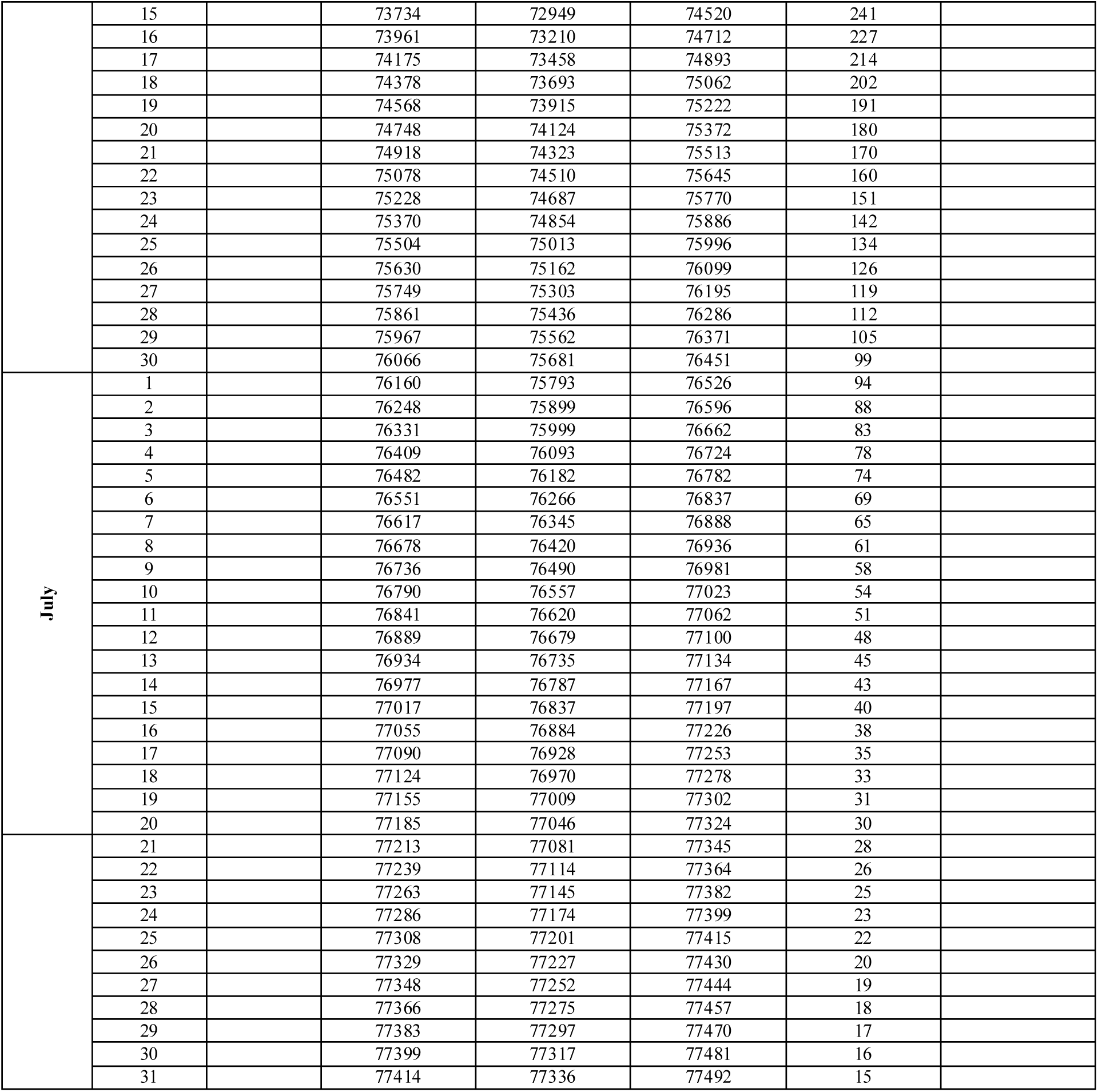
**Appendix**

